# Rapidly Adaptable Multiplexed Yeast Surface Display Serological Assay for Immune Escape Screening of SARS-CoV-2 Variants

**DOI:** 10.1101/2023.02.17.23286074

**Authors:** Joanan Lopez-Morales, Rosario Vanella, Tamara Utzinger, Valentin Schittny, Julia Hirsiger, Michael Osthoff, Christoph Berger, Yakir Guri, Michael A. Nash

## Abstract

With numerous variations in the Spike protein, including concentrated mutations in the receptor-binding domain (RBD), the SARS-CoV-2 Omicron variant significantly shifted in the trajectory of the COVID-19 pandemic. To understand individual patient risk profiles in the face of rapidly emerging variants, there is an interest in sensitive serological tests capable of analyzing patient IgG response to multiple variants in parallel. Here, we present a serological test based on yeast surface display and serum biopanning that characterizes immune profiles against SARS-CoV-2 RBD variants. We used this yeast-based multi-variant serology method to examine IgG titers from 30 serum samples derived from COVID-19-convalescent and vaccinated individuals in Switzerland and assessed the relative affinity of polyclonal serum IgG for Wuhan (B lineage), Delta (B.1.617.2 lineage), and Omicron (B.1.1.529 lineage) RBD domains. We validated and benchmarked our system against a commercial lateral flow assay and showed strong concordance. Our assay demonstrates that serum IgGs from patients recovered from severe COVID-19 between March-June 2021 bound tightly to both original Wuhan and Delta RBD variants, but became indistinguishable from background when assayed against Omicron, representing an affinity loss of >10-20 fold. Our yeast immunoassay is easily tailored and parallelized with newly emerging RBD variants.

## Introduction

The coronavirus disease 2019 (COVID-19) pandemic caused by severe acute respiratory syndrome coronavirus 2 (SARS-CoV-2) triggered a global health crisis with severe socio-economic impacts^1^. The virion of this RNA+ virus possesses four structural proteins, with the most relevant for cell entry being the trimeric Spike (S) glycoprotein. The spike protein consists of an upper globular domain (S2 domain) and a lower domain (S1) responsible for the fusion of the lipid bilayers of the virus and the host cell (S1 domain)^2–4^. The region of the S1 domain comprising amino acids N331 to T531 is the receptor-binding domain (RBD), which binds to the surface protein angiotensin-converting enzyme 2 (ACE2), mediating the entry of SARS-CoV-2 into ACE2-expressing human cells^5,6^. RBD represents a highly variable region of the viral genome, and computational analyses coupled with deep mutagenic scanning have classified therapeutic antibodies based on their ability to bind RBD epitopes and block viral entry or suffer from immune escape due to emergent mutations^7–10^. These prior studies highlight the important role of RBD in viral fitness and how it is a crucial player in processes such as infection, variant evolution, and viral interaction with the human immune system.

Diagnosing acute SARS-CoV-2 infections relies on viral RNA detection by reverse-transcription loop-mediated isothermal amplification (RT-LAMP) or reverse-transcription polymerase chain reaction (RT-PCR), which is the most used SARS-CoV-2 detection method. Conversely, serological tests detect prior infection by measuring serum antibodies against SARS-CoV-2. Serological assays typically measure anti-spike (S) or anti-nucleoprotein (N) antibodies^5^, which are generated following seroconversion of convalescent COVID-19 patients 6 to 15 days after symptom onset. These antibodies remain detectable in the blood for an extended period of time^11–13^. Consequently, serological tests are a fundamental tool to profile a patient’s immunoglobulin G antibodies (IgGs) at different time points. However, intrinsic biological titer variability and interfering effects from sample matrices make serology tests challenging to standardize and quantify^14^.

Conventional enzyme-linked immunosorbent assays (ELISA) and electrochemiluminescent alternatives are indirect tests that detect total IgG using immobilized SARS-CoV-2 antigen molecules on microtiter plates. Such assays are considered a gold standard for clinical serology^15^, and although highly sensitive, are not available for public home usage and are poorly scalable due to reagent costs, time, and personnel and equipment requirements^16^.

To address these limitations, many different serological assays have been developed. Lateral flow immunoassays (LFAs) are available for detecting IgG binding to a small number of viral antigens, including S, RBD, or N proteins. While these are rapid and low cost, they do not quantify relative or absolute antibody titers and may suffer from low sensitivity and specificity, with false-negative rates reported up to ∼15%^17^.

Micro- and nanofluidic approaches can potentially provide sensitive and specific testing at a low cost. These miniaturized assays typically use microliter sample volumes and are adaptable for multi-antigen-testing for detecting SARS-CoV-2 and influenza A antibodies in parallel^18^, as well as performing antibody affinity profiling and assessing antibody titers^19^. Miniaturized serology tests have also been built using point-of-care diabetic glucometers^14^.

However, each assay measurement scheme has associated limitations, for example, the need for specialized equipment and facilities for nanofabrication and requirements for highly trained personnel. In many assay schemes, the requirement of recombinant protein antigens as bait molecules on device surfaces is limiting in the face of rapidly emerging new variants. There remains a need to develop improved methods for SARS-CoV-2 serological testing. To overcome current limitations in diagnostic serology of COVID-19, an ideal platform would be scalable to the population level, rapidly adaptable to newly emerging variants, highly sensitive, and highly specific at a low cost per test.

Yeast surface display is an intriguing biotechnology platform for developing novel serology assays. Plasmids encoding the protein to be displayed fused with a yeast mating factor (Aga2) are transformed into yeasts and induced for gene expression, providing a robust cell line capable of displaying the protein of interest on the outer cell wall of the yeasts. Since no recombinant protein purification is required, the displayed protein can be rapidly exchanged through a simple cloning and transformation procedure following the emergence of new viral variants. With its eukaryotic protein quality control system and posttranslational modification machinery, yeasts can display complex proteins, such as SARS-CoV-2 surface and structural proteins. Indeed, yeast surface display has allowed a deep understanding of SARS-CoV-2 infection evolution and immune escape of therapeutic antibodies^8–10,20,21^. We hypothesized that yeast surface display can offer a unique and scalable platform on which to base serological assays.

Here we developed a workflow for the detection of antibodies (i.e. IgGs) specific for SARS-CoV-2 RBD variants displayed on yeast cells with high sensitivity and specificity. We validated our yeast serological assay for SARS-CoV-2 and report seroprevalence and antibody profiling in a small cohort of convalescent or SARS-CoV2-naive mRNA vaccinated subjects. Our assay revealed that IgGs from mRNA-vaccinated individuals as well as individuals recovered from severe infection between March and June 2021 strongly recognize original and delta RBD variants but suffered a 10-20 fold loss in apparent affinity when assayed against the RBD of the Omicron variant. This validates other studies and helps explain the higher prevalence of breakthrough infections observed with Omicron throughout 2022.

## Methods and Materials

### Chemicals and antibodies

Antibodies: 6x-His Tag Monoclonal Antibody (MA1-21315); Goat anti-Human IgG (H+L) Cross-Adsorbed Secondary Antibody, Alexa Fluor 594 (A-11014); Goat anti-Mouse IgG (H+L) Cross-Adsorbed Secondary Antibody, Alexa Fluor 594 (A-11005); SARS-CoV-2 Spike Protein (RBD) Recombinant Human Monoclonal Antibody (T01KHu, 703958); 6x-His Tag Monoclonal Antibody-DyLight 488 (MA1-21315-D488) were purchased from Invitrogen-Thermo Scientific. Human recombinant ACE2 Protein (hACE2-hFc Tag, 10108-H02H) was purchased from SinoBiological Europe GmbH, Germany. The SARS-CoV-2 rapid antibody test (9901-NCOV-02C) was purchased from Roche, Switzerland.

### Yeast Display of SARS-CoV-2 proteins

Three different receptor binding domains (RBD) constructs belonging to the most frequent spike protein S1 from SARS-CoV-2 variants of concern were designed by extracting the phylogenetic sequences of RBD variants from the Nextstrain platform (https://nextstrain.org/ncov/gisaid/global) and employing codon-optimization for *Saccharomyces cerevisiae*. The DNA sequences were synthesized by Twist Biosciences. EBY100 yeasts carrying pYD1 plasmids with an empty cassette negative control (EC), RBD sequences from Wuhan (B lineage), Delta (B.1.617.2 lineage), or Omicron (B.1.1.529 lineage) SARS-CoV-2 were plated on yeast synthetic drop-out medium (YS)–Trp + 2% (w/v) Glucose (Gluc) + 100 μg/mL Ampicillin (Amp) plates. Precultures were grown overnight at 30°C, 180 rpm in YS–Trp+2% (w/v) Gluc + Amp until saturation. The next day, the cultures were diluted to OD_600_=0.4 in YS–Trp + 1.8% (w/v) Galactose (Gal) + 0.2% (w/v) Gluc + 100 μg/mL Amp + Citrate/Phosphate buffer pH 7.0 and cultured at 20°C, 180 rpm for 48 h. After the induction period, cells were spun down, washed with ultrapure water, and resuspended in PBS + 0.5% BSA + 0.5 mM EDTA pH 7.5 buffer.

### Flow cytometry

2×10^6^ cells from induced cultures were collected, washed with 1 mL PBS + 0.5% BSA + 0.5mM EDTA pH 7.5 buffer, and pelleted. The cell pellets were labeled at the C-terminus using a primary mouse anti-His6 mAb (1:500) for 30 min at room temperature. After incubation, cells were rinsed with 1 mL PBS-BSA-EDTA buffer and pelleted again. As secondary labeling, cells were labeled with a goat anti-mouse mAb conjugated to AlexaFluor 488 dye (1:500) for 20 min at 4°C, protected from light. After incubation, cells were washed five times and resuspended in PBS+0.1% BSA for flow cytometry analysis. Full-length protein display was assessed by flow cytometry on an Attune NxT flow cytometer (Thermo Scientific, USA) equipped with a high-throughput autosampler and 488 and 561 nm lasers, using Attune Nxt Software v3.2.1 (Life Technologies) or a FACSMelody sorter (Becton Dickinson) and analyzed with FlowJo 10.8.1 (Becton Dickinson). The fraction of cells displaying the protein of interest was obtained by gating out the uninduced population. The relative protein amount per cell was observed as the median fluorescence of the analyzed single cells displaying population.

### Swiss cohort of convalescent and vaccinated individuals

Plasma samples from twenty non-vaccinated individuals treated for severe COVID-19 and discharged from the intensive care unit (ICU) at the University Hospital of the University of Basel were collected in early 2021. At enrolment, written informed consent was collected. In addition, ten serum samples from healthy donors vaccinated against SARS-CoV-2 were collected at the University Hospital, and the serological status of the participants was recorded (**Table 2**). Sampling was performed in accordance with the Declaration of Helsinki. The donors included adults of both sexes. This study was approved by the Ethical Committee of Norwest-Central Switzerland (EKNZ) in Basel (Project-ID 2021-00214).

**Table 1.** Lineage description of displayed SARS-CoV-2 RBDs from VOCs in Switzerland.

| Variant | Lineage | Amino acid mutations |
| --- | --- | --- |
| RBD WT | Wuhan | Reference |
| RBD Delta | B.1.617.2 | L452R, T478K |
| RBD Omicron | B.1.1.529 | G339D, S371L, S373P, S375F, K417N, N440K, G446S, S477N, T478K, E484A, Q493K, G496S, Q498R, N501Y, Y505H |

**Table 2.** Patient cohort and employed sera.

|  | Features | n<br>(female/male) | Mean age<br>(range) | Date of<br>acquisition | Sample<br>acquisition |
| --- | --- | --- | --- | --- | --- |
| Patients | Severe symptoms, hospitalized (ICU), unvaccinated | 20 (7/13) | 55(25-76) | March & April 2021 | 15-20 days after symptom onset (24 h after ICU discharge) |
| Vaccinated individuals | 2xmRNA-1273 (Spikevax®, Moderna) | 10 (5/5) | 33 (27-43) | December 2021 | 28 days post 2nd Covid Vaccination |

### Serum Depletion

To reduce the non-specific binding background, serum samples were depleted by incubating 150 μL of serum with 400×10^6^ induced empty cassette (negative control) yeast cells for 3 h at 20°C and 1400 rpm. After incubation and cell pelleting, the serum supernatant was transferred to a tube with the same number of empty cassette cells. This second incubation was continued as described before for a total of 6 h. Finally, the serum was recovered and filtered with a 0.45 μm-pore microcolumn and stored at -80°C.

### Serology of patient sera by yeast display

For serum titration on yeast, sera were diluted over a range from 1:25 to 1:1.5×10^6^ in 96-well microtiter U-bottomed plates (351177, Falcon) containing prewashed 2.0×10^4^ cells displaying either the negative control or one of the three different RBD variants mixed with 8.0×10^4^ wild-type EBY100 non-induced yeast cells. The cells were prepared in stocks at the correct ratio and seeded into the wells. After buffer removal, cells in each well were resuspended in 30 μL of the corresponding depleted-serum dilution, and the plate was incubated for 1 h at 4°C and 1000 rpm. The plate was then washed twice with ice-cold PBS-BSA-EDTA buffer. IgGs in the sera were detected using a goat-anti-hIgG-Alexafluor-594 mAb, and the displayed construct was simultaneously detected using a goat-anti-6xHis-DyLight-488 mAb (30 μL total, 1:500 both mAbs) in PBS+BSA buffer. After 30 min of incubation at 4°C and 1000 rpm, the cells were washed once and resuspended in ice-cold 150 μL of PBS-BSA-EDTA buffer. Flow cytometric analysis of double-labeled single cells was performed as previously described. The fraction of cells displaying the protein of interest was obtained by gating out the uninduced cell population. From the displaying cell population, the fraction of cells bound to sera antibodies (percentage of cells) was recorded. Python together with Matplotlib.pyplot, Pandas, and NumPy packages were used to plot and fit the data to the Hill model. EC_50_ values (i.e. the dilution factor required for half saturation of binding) for each construct per serum were obtained from the fit. Significance was calculated by pairwise non-parametric multiple comparison ANOVA (Friedman’s test) using the internal negative control’s (empty cassette) EC_50_ values as the reference value for all three RBD variants’ EC_50_s.

## Results & Discussion

### Design and test development

We designed our yeast immunoassay for the detection of serum antibodies against a negative control and three RBD constructs belonging to frequent SARS-CoV-2 variants of concern (VOCs) across Switzerland (WT (Wuhan), Delta, and Omicron) (**Fig. S1 and Table 1**). Yeast surface display provides several advantageous features for our purposes, including eukaryotic protein production and post-translational modifications, facilitation of correct folding, ease of labeling of bound/displayed constructs, robust handling, ease of genetic manipulation, and a capacity for upscaling^22^.

The serological assay simultaneously employs four yeast cell lines to pan human sera against RBD variants. Yeast displayed RBDs were incubated with serial dilutions of patients’ sera followed by immunolabeling and detection of both the level of displayed RBD variant (green fluorescent channel) and bound immunoglobulin (red fluorescent channel) using analytical flow cytometry **(Fig. 1a**). The bound fraction’s values per RBD variant were fitted to a 4-parameter logistic model to obtain a relative titer of antiRBD-IgG, expressed as an EC_50_ value (**Fig. 1b**).

**Figure 1.**
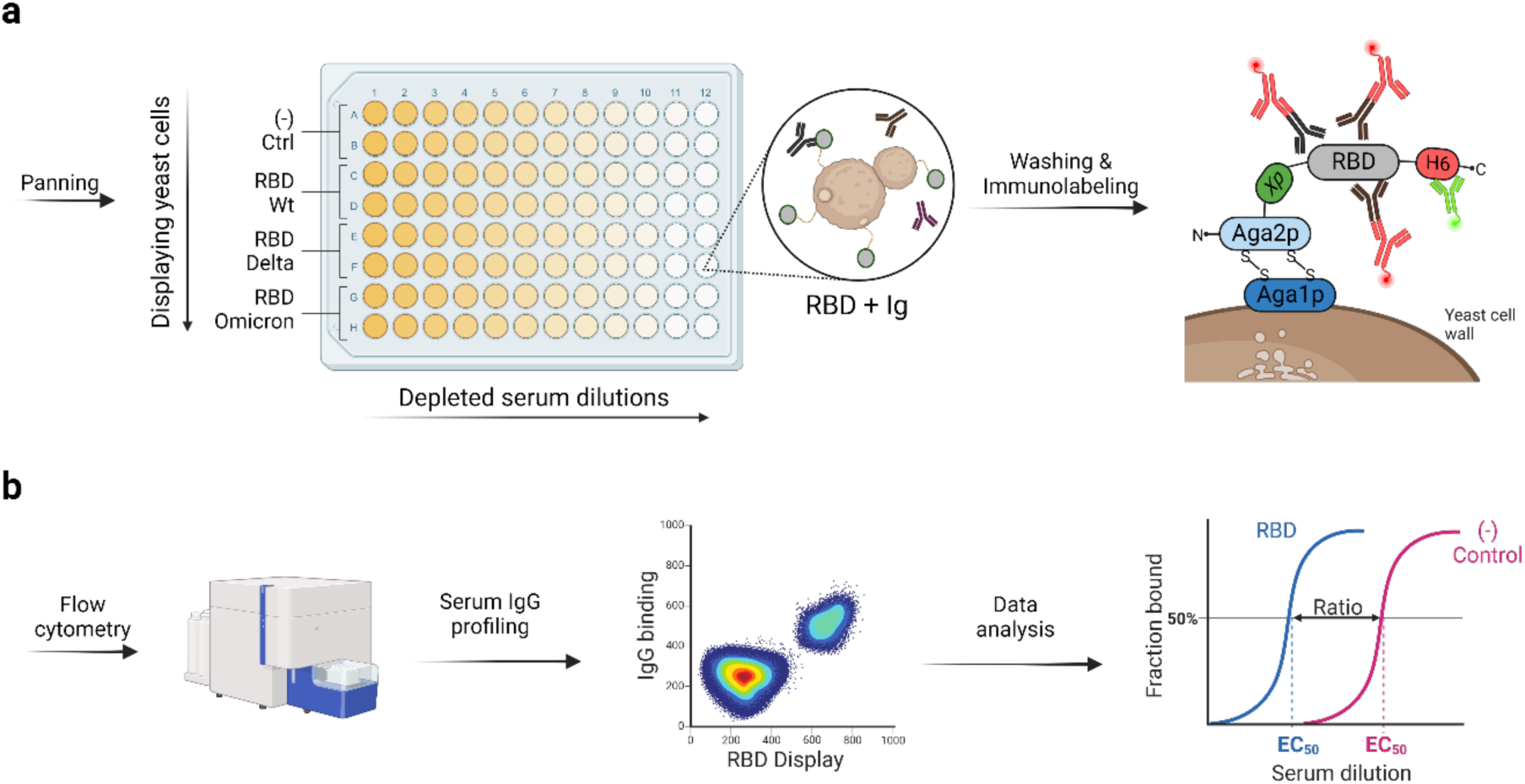
Overview of the yeast serological test. **a**. Dilution series of patient sera preprocessed by depletion with empty cassette yeast cells were exposed to yeasts displaying specific RBD variants in multititer plates in order to titrate binding of RBD-specific human IgGs. One-step immunostaining allowed the quantification of cells stained double positive for correct display (shown as green fluorescent mAbs) and specific binding to RBD (shown as red fluorescent mAbs). **b**. High-throughput analytical flow cytometry of samples in duplicate quantified IgG binding to each VOC as compared to an empty cassette negative control yeast.

#### Evaluation of RBD binding to ACE2 using yeast display

Several studies have successfully employed yeast display of SARS-CoV-2 RBDs and related coronaviruses to map mutations that lead to immune escape^8–10,21,23,24^. Supported by these previous reports, we optimized yeast surface display of RBD variants and evaluated binding to human ACE2 (ACE2)^25–29^. We found the three RBD constructs were optimally displayed following 1.8% (w/v) galactose (Gal) + 0.2% (w/v) glucose induction at 20°C, pH 7.0 for 48 h. After expression, cells displaying the three RBD variants were incubated with fluorescently labeled ACE2. Flow cytometry analysis confirmed that the yeast-expressed RBDs specifically bound ACE2 (**Fig. S2**).

### Assay optimization

#### Minimization of ligand depletion effects

When assessing the relative affinity of ligands to their cognate binders immobilized on a cell or solid surface, one artefact that can lead to overestimation of the dissociation constant is referred to as the ligand depletion effect. If the soluble ligand (in this case, serum IgGs of unknown concentration) is not in significant excess to the number of binding sites on the cell surface, affinity titrations will mistakenly report higher dissociation constants due to depletion of the free ligand.^30–32^ For many yeast-based binding assays, the sample volumes can be scaled up at a fixed cell number, however, since collected patient sample volumes were constrained, we chose to decrease the number of cell surface binding sites by reducing the number of displaying cells present in the reaction. Both approaches (increasing reaction volume or decreasing binding sites at the cell surface) lead to a higher ratio of soluble IgG to displayed RBD, which allows the approximation of free ligand concentration with total ligand concentration, simplifying experiments.^33,34^

We fixed the consumed sample volume per pair of titration curves to 15 μL to allow testing samples in duplicate against three RBD variants plus a negative control in parallel. Given this maximum serum volume, we evaluated the effect of decreasing RBD displaying cell numbers in the reaction mixture to find conditions that avoided ligand depletion effects of greater than 10% for each tested dilution. We decreased the number of RBD displaying yeasts while increasing the number of non-displaying yeast cells at a fixed total cell number per reaction of 100,000 cells, which was required for cell pelleting and flow cytometry analysis.

Four ratios of RBD-displaying:non-displaying yeast cells were tested and incubated in duplicate with 30 μL of serial dilutions of serum ranging from 1:25 to 1:1.5×10^6^ (**Table 3**). Cells were then stained for RBD display and IgG binding and analyzed by flow cytometry (**Fig. 2**). As expected, smaller ratios of displaying to non-displaying cells resulted in left-shifting of the titration curve. The ratio of 20,000 RBD-displaying cells: 80,000 non-displaying cells was chosen for all further experiments. At ≤20,000 RBD displaying cells, there were no ligand depletion effects observed, the calculated EC_50_ converged, and enabled correct measurements of binding events (data for smaller ratios are not shown for clarity). These control tests demonstrate that the titration curves produce the lowest EC_50_ values for more highly diluted numbers of RBD-display cells. This is an important consideration for conducting our assays in a regime that avoids ligand-depletion effects^33^.

**Table 3.** Drop-in screening for reducing ligand depletion effects in binding experiments.

| Variant RBD | Induced cells | Non-induced cells | Induced fraction | $EC_{50}$ | $\sigma EC_{50}^*$ |
| --- | --- | --- | --- | --- | --- |
| WT | $1.0 \times 10^5$ | 0.0 | 100% | $1.432 \times 10^{-3}$ | $2.22 \times 10^{-4}$ |
| WT | $5.0 \times 10^4$ | $5.0 \times 10^4$ | 50% | $5.51 \times 10^{-4}$ | $2.9 \times 10^{-5}$ |
| WT | $2.0 \times 10^4$ | $8.0 \times 10^4$ | 20% | $3.99 \times 10^{-4}$ | $2.4 \times 10^{-5}$ |
| Neg. Ctrl | $1.0 \times 10^5$ | 0.0 | 0% | $3.01 \times 10^{-3}$ | $2.74 \times 10^{-4}$ |

\* $\sigma EC_{50}$ : standard deviation of $EC_{50}$ duplicates.

**Figure 2.**
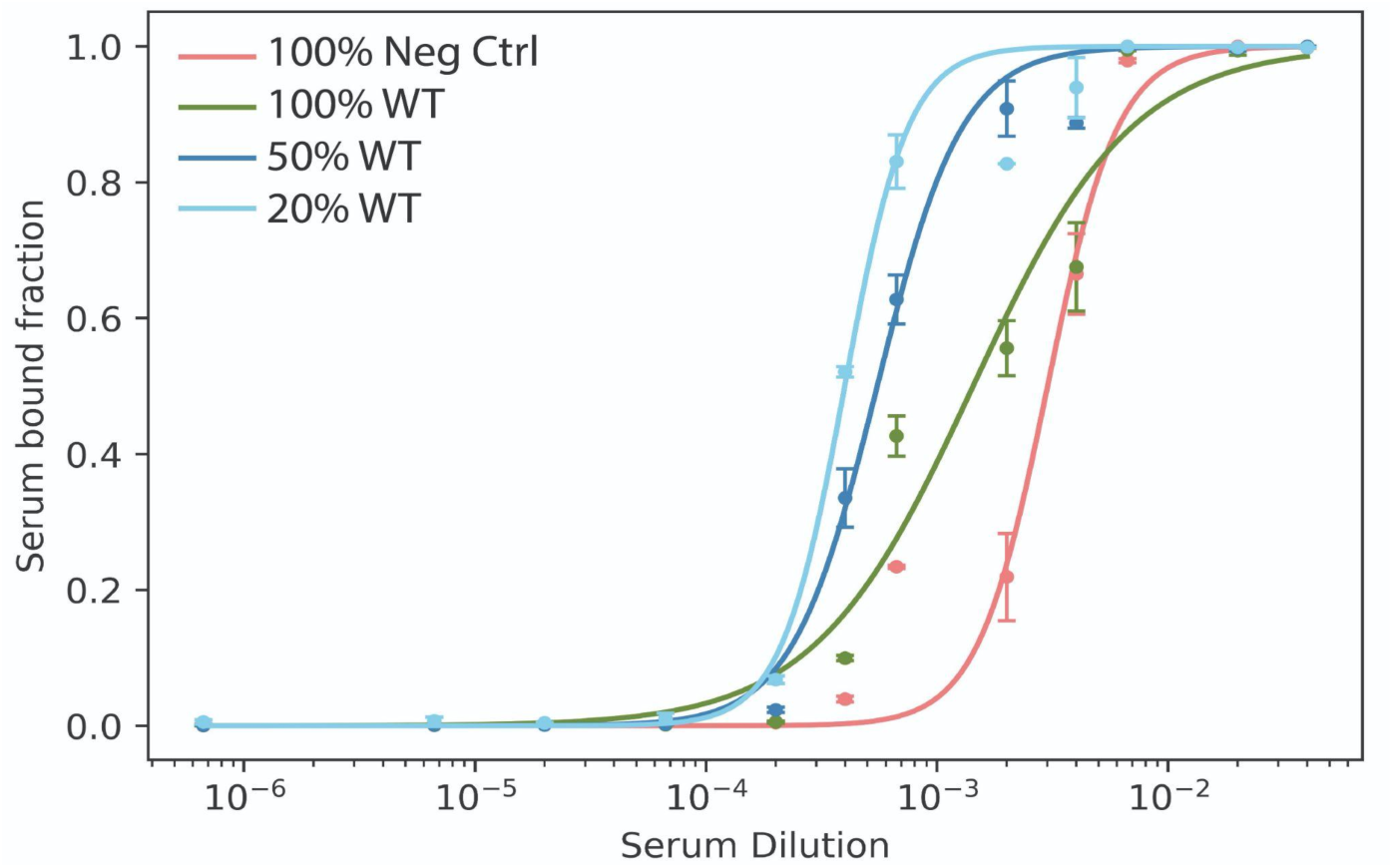
Minimizing ligand depletion effects. Yeast cells displaying WT-RBD or empty cassette were mixed in different ratios with non-induced cells to reduce the number of receptors on the yeast cell wall and minimize ligand depletion artefacts. Percentages refer to the fraction of displaying cells of each cell type. Due to ligand depletion effects, at high numbers of WT-RBD displaying cells, the binding curve was shifted further to the right and overestimated EC_50_. Neg ctrl: a displaying cassette that included Aga2-(Xpress tag)-(6x Histidine tag) but no RBD used as negative control; WT: Wuhan reference RBD.

#### Reduction of non-specific binding by serum-depletion negative selection

One major challenge of panning yeast cells directly against patient serum is the non-specific binding of sample IgGs to the yeast surface. This interaction sets the assay background against which a specific signal must be detected. To address this issue, we depleted serum samples of immunoglobulins that non-specifically bound the yeast surface prior to performing the SARS-CoV2-RBD binding assay. Sera were incubated with yeast cells displaying an empty cassette construct that displayed only the Xpress and 6x histidine tags but lacked the RBD construct. Optimization of this step involved testing different variables, such as incubation time, number of cells, number of depletion steps, filtration steps, and presence or absence of the displaying cassette. Finally, the benefits of serum depletion were validated on positive serum for anti-SARS-CoV-2 antibodies (pre-Omicron wave). Cells displaying RBD constructs and the negative control were incubated with serial dilutions of depleted and non-depleted serum, stained as previously described, and analyzed by flow cytometry. **Figure 3** shows a clear difference in binding profiles between depleted the non-depleted serum. Non-depleted serum showed high non-specific IgG binding to the cells and resulted in high background signals (**Fig. 3a**). In the case of the depleted serum, the curves of the WT and Delta RBDs showed significantly higher binding than the negative control and omicron RBD (**Fig. 3b**). No binding was observed for the omicron variant which generated signal in the same dilution range as the negative control. Ultimately, two incubation steps of 3 h with 400 million cells displaying the negative control empty cassette were used for serum depletion conditions to reduce non-specific signals.

**Figure 3.**
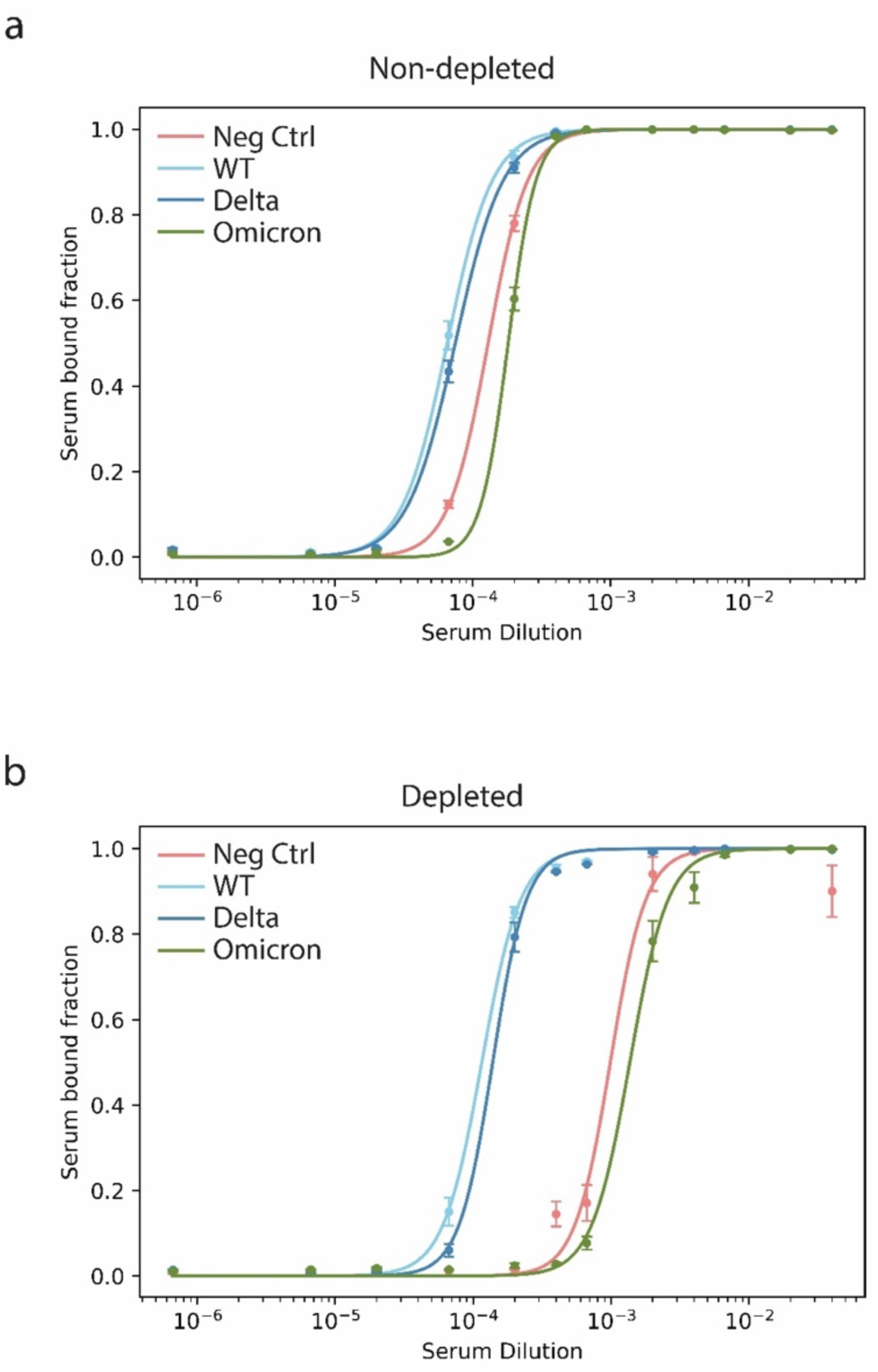
Effect of serum depletion of non-specific binders. Human sera were depleted of antibodies that non-specifically adhered to yeasts by incubation with yeasts displaying an RBD-negative empty cassette (in this case, Aga2-Xpress tag-6x His tag, labeled negative control). Pre-incubation of the sera with 400 million cells displaying the empty cassette followed by filtration decreased non-specific binding of serum proteins to yeast cells. The employed serum was validated positive for SARS-CoV-2 by RT-PCR and for anti-SARS-CoV-2 antibodies (pre-Omicron wave) by lateral flow assay. **a**. High non-specific binding masks the true binding signal and confounds detection of RBD-specific IgGs in samples that were not pre-depleted. **b**. The effect of serum depletion enhanced the true binding signal, revealing the true binders from the non-specific. Error bars: standard error of the mean (s.e.m.) from n = 2 independent experiments. Neg ctrl: yeast display cassette that includes Aga2-Xpress tag-6x His tag.

### Immunoprofiling of convalescent sera

The control experiments to validate depletion of non-specific binding IgGs and assess ligand depletion effects gave us confidence that the assay can specifically detect IgG-binding to RBD-yeast cells panned against sera. To characterize the presence of antibodies against SARS-CoV-2, sera from the clinical study were first tested with a Roche SARS-CoV-2 IgG/IgM rapid antibody lateral flow test with a sensitivity of 99.03% and a specificity of 98.65%, as reported by the manufacturer. Of the twenty convalescent sera tested, 20% showed a double IgG/IgM negative result on the lateral flow test (**Table 4, Fig S3**). This result may be attributable to these patients being immunosuppressed and non- or only very weakly seroconverted. All sera of vaccinated individuals tested with the SARS-CoV-2 rapid antibody lateral flow were positive (**Fig. S4**).

**Table 4.** Lateral flow assay of human sera. Convalescent patient (not shaded), vaccinated (shaded in grey) and commercially validated SARS-CoV-2-negative (shaded in blue) human sera were analyzed with a Roche SARS-CoV-2 rapid antibody test.

| Serum sample | IgG | IgM | Serum sample | IgG | IgM | Serum sample | IgG | IgM |
| --- | --- | --- | --- | --- | --- | --- | --- | --- |
| (-) Control | - | - | 15 | + | + | 31 | - | - |
| (+) Control | + | - | 16 | ++ | + | 32 | - | - |
| 1 | - | - | 17 | - | + | 33 | - | - |
| 2 | + | - | 18 | ++ | + | 34 | - | - |
| 3 | ++ | + | 19 | ++ | + | 35 | - | - |
| 4 | + | + | 20 | ++ | + |  |  |  |
| 5 | - | - | 21 | ++ | - |  |  |  |
| 6 | + | + | 22 | ++ | - |  |  |  |
| 7 | ++ | + | 23 | ++ | - |  |  |  |
| 8 | ++ | + | 24 | ++ | - |  |  |  |
| 9 | - | - | 25 | ++ | - |  |  |  |
| 10 | + | + | 26 | ++ | - |  |  |  |
| 11 | + | - | 27 | ++ | - |  |  |  |
| 12 | - | - | 28 | ++ | - |  |  |  |
| 13 | ++ | + | 29 | ++ | - |  |  |  |
| 14 | - | + | 30 | ++ | - |  |  |  |

### Multiplexed serology of anti-SARS-CoV-2 IgG from human sera

We next tested the binding of yeast displayed RBD variants to serial dilutions of twenty pre-processed sera of COVID-ICU discharged individuals and 10 pre-processed sera of vaccinated individuals using the optimized assay conditions described above (**Table S3, Fig. S5**, and **S6**). The serology results were interpreted based on shifts of the titration curve midpoint (EC_50_) for different RBD variants compared to the negative (empty cassette) control yeast. A leftward shift of the titration midpoint (toward higher dilution factors) indicated binding of serum IgGs above background. A lower EC_50_ value of a serum sample against a particular variant therefore indicated a higher titer and/or tighter binding of serum IgGs specific to that variant.

We estimated the limit of detection of the yeast assay in terms of serum dilution factor by calculating an average of the three highest dilutions still detectable from 3 validated positive sera and found it to be 1:1×10^−6^. To quantify the sensitivity of our test, we used a validated positive serum with the lowest anti-SARS-CoV-2 IgG titer. In comparison, we analyzed five commercially available single-donor human SARS-Cov-2 Negative Sera (ISERSCOV2N100UL, Innovative Research, USA) to test the specificity of the yeast serological assay. A verified positive serum was employed to validate the RBD strains and labeling reagents for the specificity test. The five negative commercial sera tested negative in our yeast immunoassay (**Table S3, Fig. S7, and S8**). We concluded that our yeast immunoassay correctly distinguished the negative samples from the positive one.

The values of EC_50-neg_ and EC_50-RBD_ were employed to calculate the quotient EC_50-neg_/EC_50-RBD_ of 3.25, which we defined as the threshold at which to classify a sample as positive. In the case of observing little to no shift in the binding curve relative to the negative control (i.e. EC_50-neg_/EC_50-RBD_ < 3.25), the serum was classified as negative for anti-RBD IgGs. In the case of a serum lacking IgGs for a specific RBD VOC, no shift was observed in the corresponding variant curve, which was labeled as negative for that variant (**Fig. 4a**).

**Figure 4.**
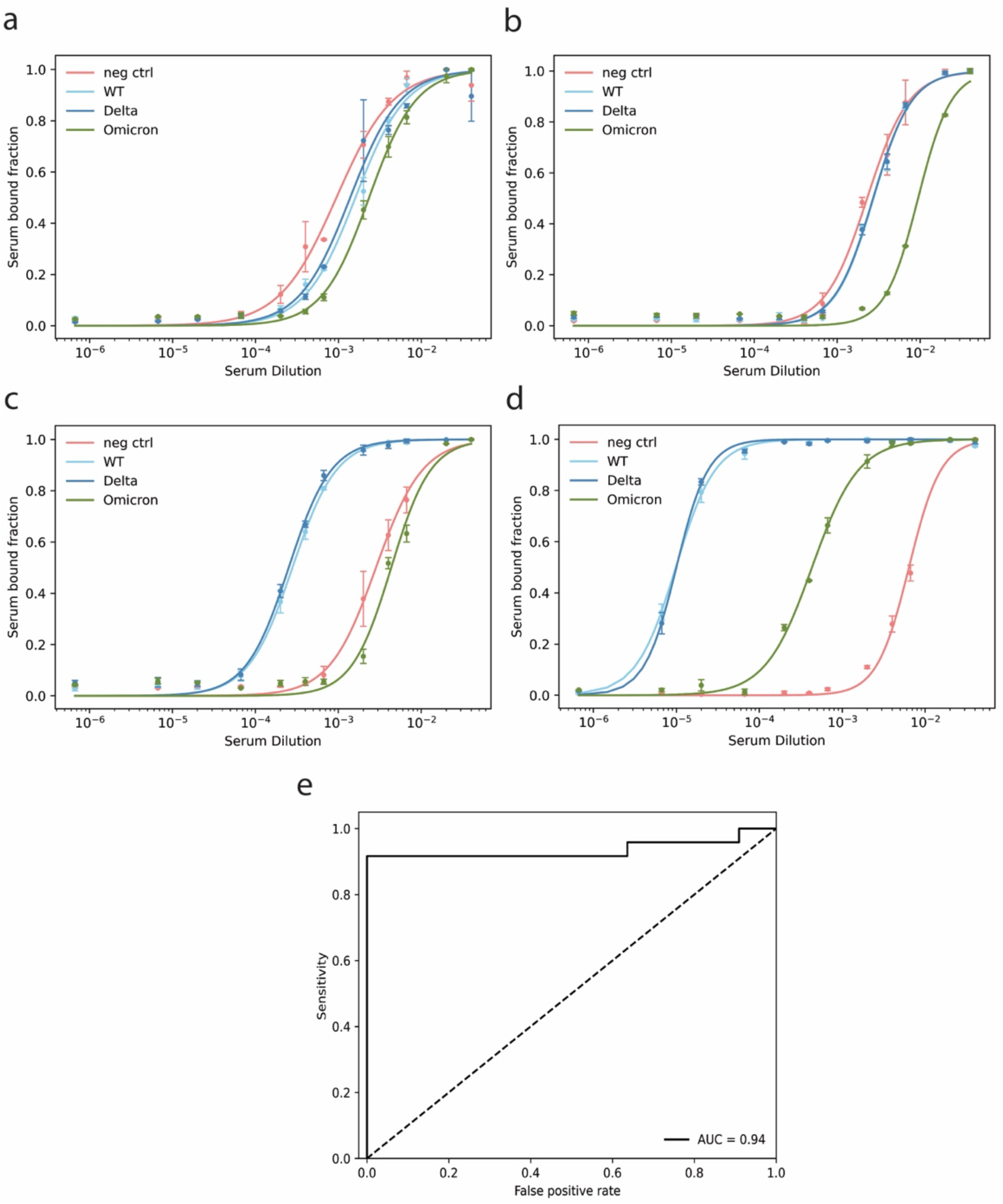
Representative yeast serological test results. **a**. Typical test results of a validated negative serum. The sample EC_50_ values spanned the same value range as the negative control (EC_50-neg_/EC_50-RBD_ < 3.25), indicating that the serum was negative for anti-RBD IgGs for all variants. **b**. Plot showing IgG isotype specificity effect. The test was designed to identify potential anti-RBD IgGs. If the serum sample contained only IgM or IgG/IgM against other antigenic proteins of the virion (e.g., N or full length S epitopes) but not against RBD, it is expected to appear negative on the yeast assay but positive on the Roche lateral flow IgM test line. **c**. Typical test results of a validated positive serum (EC_50-neg_/EC_50-RBD_ > 3.25). The yeast assay was positive for IgGs against WT and Delta RBD, but not Omicron. **d**. Test results from a sample obtained from a vaccinated individual. The yeast assay showed that the vaccinated serum had a high titer of IgGs recognizing WT and Delta RBD, and lower titer for IgGs recognizing Omicron RBD. Error bars represent the standard error of the mean (s.e.m.) from n = 2 independent experiments. **e**. The receiver operating characteristic curve shows the true positive (sensitivity)/false positive (1-specificity) fraction ratio of the yeast serological assay relative to the reference Roche rapid antibody test. AUC: Area under the curve.

The results showed a specificity of 100% and a sensitivity of 84.6% for SARS-CoV-2 WT when taking the Roche rapid antibody LFA test as ground truth. We assessed the possible causes for the modest sensitivity value. The four false negatives were divided into two groups. The first group originated from sera that tested positive for IgGs but negative for IgMs by the Roche lateral flow test. One possible explanation for the discordant results is that the Roche lateral flow test used whole spike and nucleocapsid proteins as bait antigens for antibody detection, while our yeast assay only uses RBD (**Fig. 4b**). Sample matrix interferences or low IgG titer could explain these false negatives.

The second group of false-negatives comprised sera that tested positive for IgM only by the Roche lateral flow test. In the yeast assay, the same sera were classified as negative because the anti-Human IgG secondary antibody that we used only detects IgGs (not IgMs) (**Fig. 4b**). Immune responses based only on IgMs tend to be weaker and less neutralizing as compared to IgG responses^35^. The yeast serological assay correctly classified the remainder of the sera as true positives, and typical positive curves showed a significant shift to higher dilutions than negative controls (**Fig. 4c**). Our assay further allowed us to observe differences in blood antibody titers specific for different RBD variants (**Fig. 4d**). Considering that we designed the yeast serological test to identify IgGs (and not IgMs) in sera, if we take the Roche LFA IgG test line as ground truth, then the yeast assay sensitivity reached 92%. A confusion matrix showed a positive predictive value (PPV) of 100% and a negative predictive value of 83% (**Fig. S9**), emphasizing the high accuracy and confidence of the yeast immunoassay. We found that the area under the receiver operator characteristic curve (AUC) was 94%, demonstrating a significant ability of our assay to detect serum IgGs from convalescent COVID-19 patients (**Fig. 4e**). These results show the utility of our yeast serological assay in detecting anti-RBD antibodies, quantifying antibody titers, and testing for immunity to a specific variant, providing potentially useful clinical information.

These results demonstrate an ability of the yeast serological assay to provide quantitative IgG profiles on multiple RBD variants in a test format that is faster and more convenient than neutralization assays. The serological data from the yeast test, such as the presence of putative neutralizing antibodies against SARS-CoV-2, along with antibody titers, and the IgG titer specificity to a specific variant all provide an additional layer of clinically relevant information that can be utilized for a more personalized immunization strategy.

### Validation against the Elecsys anti-SARS-CoV2-IgG/M assay

Ten of the sera belonging to vaccinated individuals were further analyzed with a Roche Elecsys® Anti-SARS-CoV-2 electrochemiluminescence sandwich immunoassay to quantify the titers of IgG and IgM binding to SARS-CoV-2. The same sera were analyzed by the yeast serological immunoassay. Our results correlated well with the reported titers suggesting that the EC_50_ values obtained from the yeast serology assay can be interpreted as a semiquantitative determination of IgG titers (**Fig. 5** and **Fig. S4**).

**Figure 5.**
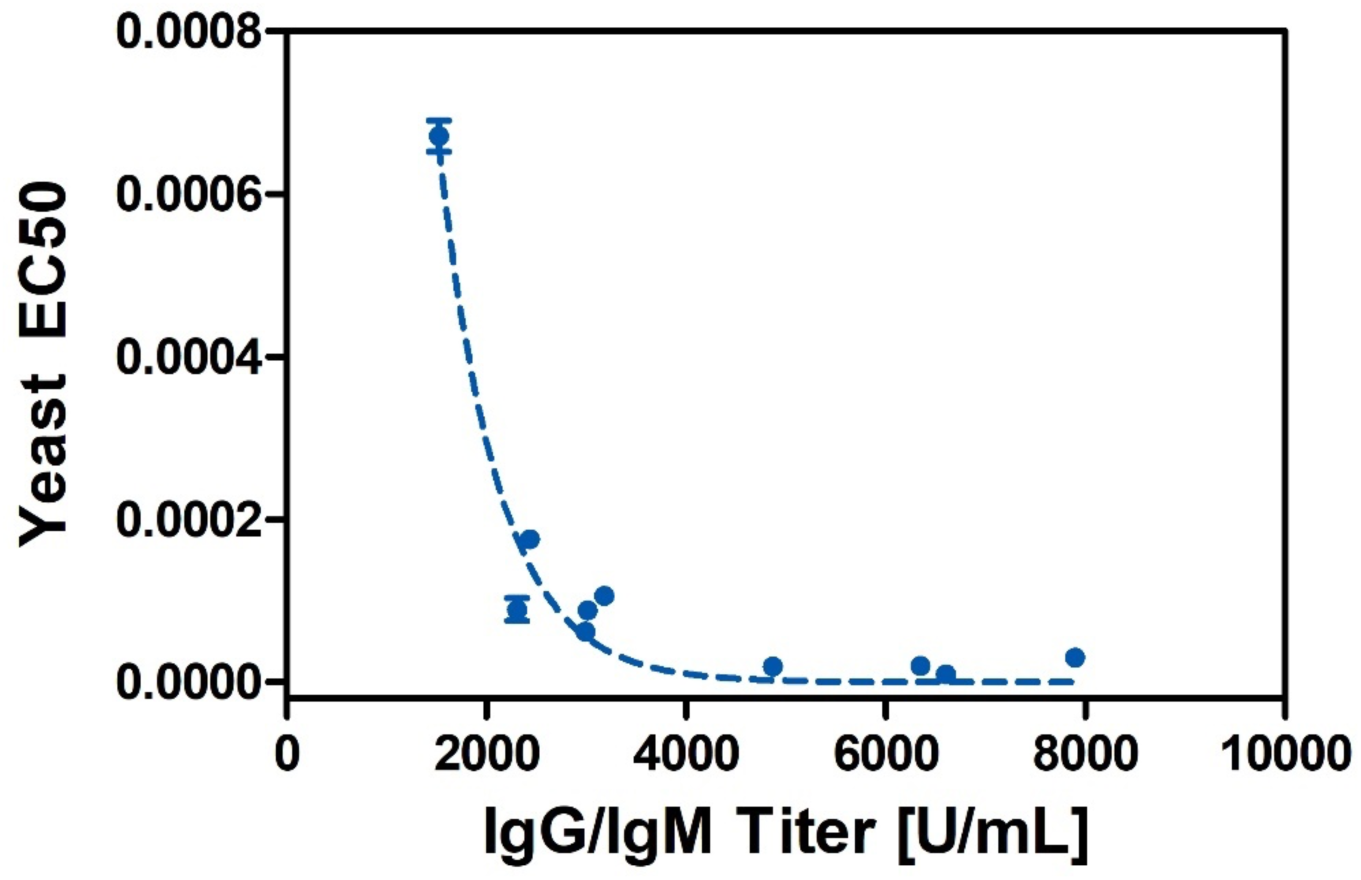
Comparison of EC_50_ values against IgG titer determined by Roche Elecsys on vaccinated subjects. Ten vaccinated sera with known IgG/IgM titers (Anti-SARS-CoV-2 (IgG/IgM) Roche Elecsys® assay) were analyzed by the yeast serological test. In the yeast assay, low EC_50_ values indicated a high IgG titer. The EC_50_ values inversely correlate with the reported titer of the serum samples determined by the commercial Elecsys electrochemiluminescence sandwich immunoassay (R^2^ = 0.9537). Error bars represent the standard error of the mean (s.e.m.) from n = 2 independent experiments.

### Yeast surface display assay detects immune escape against the Omicron variant in serum of convalescent and vaccinated subjects

Currently available antibody assays use the Wuhan WT spike or RBD protein as the antigen bait. Due to mutations in the RBD of VOCs, specifically Omicron, these tests can fall short in determining immunity against newly emerged VOCs. Omicron breakthrough infections occur at high rate even in individuals with very high anti-S-IgG against the Wuhan strain. Therefore, serological tests are needed to measure antibodies against different viral variants in parallel.

One advantage of the yeast serological assay is that it easily allows differentiation of IgG titers against different RBD variants (**Fig. 6a**). We observed no substantial differences between the convalescent sera binding to WT or Delta RBD (EC_50-Wt/Delta_ = 1×10^−4^). However, there was a significant reduction (1-2 orders of magnitude) in the binding to RBD Omicron (EC_50-Omicron_ = 5×10^−2^) (**Fig. S10a**). This result is characteristic of patients that were infected by pre-Omicron variants before the end of 2021. Immune escape of SARS-Cov-2 Omicron based on evasion of neutralizing IgGs in serum has been reported^21,36^. The observed EC_50_ values from convalescent patients confirmed the lower titer of antibodies targeting omicron’s RBD.

**Figure 6.**
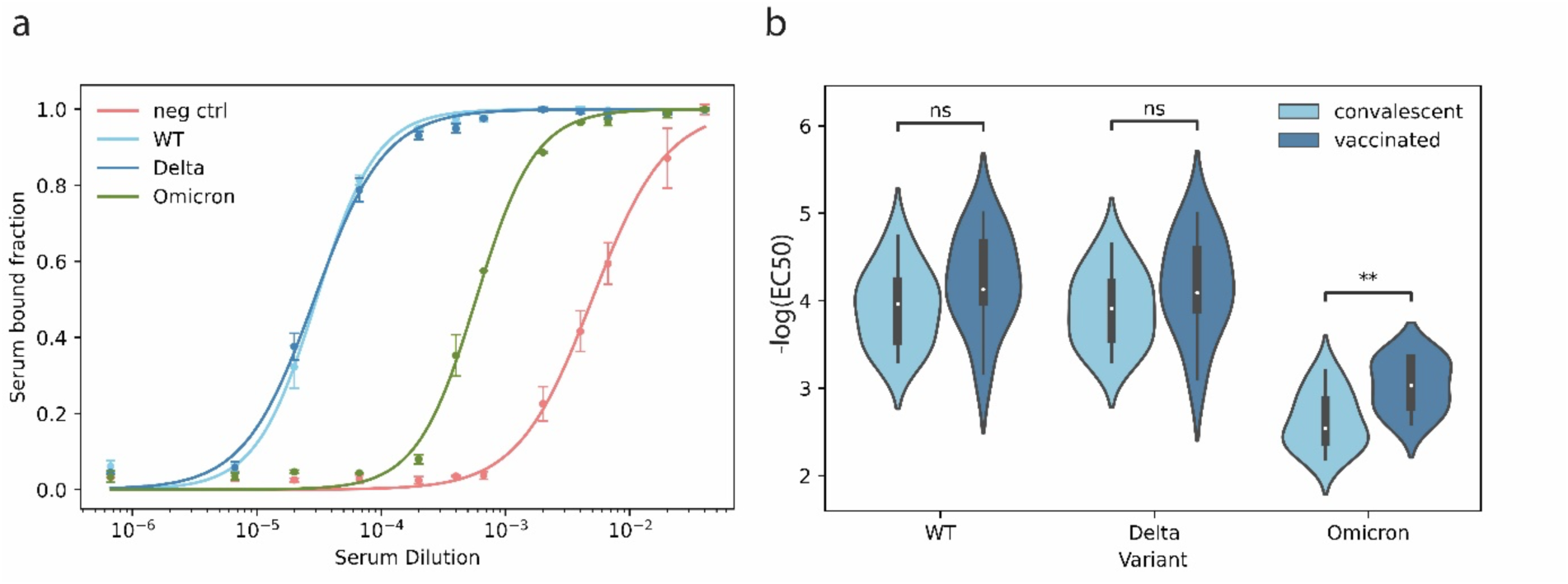
Comparative analysis of immunity between convalescent and vaccinated sera. **a**. Representative yeast assay results of vaccinated serum. The yeast serological assay revealed high IgG titer and specific recognition of WT and Delta RBD, but only moderate recognition of the Omicron RBD variant. Error bars represent the standard error of the mean (s.e.m.) from n = 2 independent experiments. **b**. Comparison of IgG titers against different RBD variants among convalescent and vaccinated sera showed the vaccination’s impact on IgG titers against SARS-CoV-2 RBD. Significance from ANOVA is shown as ns: p ≥ 1.0; **: 1.0×10_-3_ < p ≤ 1.0×10^−2^.

Among the vaccinated sera, we found that the titer of IgGs against RBD WT and Delta were statistically equivalent (EC_50-WT/Delta_ = 2×10^−4^). Conversely, the titer of IgGs to the RBD Omicron was significantly lower (EC_50-Omicron_ = 1×10^−3^)(**Fig. S10b**). The reported binding behaviors are in considerable agreement with previously described trends^36–38^ and demonstrate that omicron is characterized by significant IgG antibody escape from recovered and vaccinated individuals.

We then compared the IgG titer acquired from natural immunity after infection against the vaccination-induced IgG response. In both populations (convalescent and vaccinated), there was a significant difference between the RBD WT and Delta to the Omicron variant (20 to 100-fold). When comparing the response of two populations for each RBD VOC (**Fig. 6b**), we observed no significant difference in binding to RBD-WT and Delta variants. However, there was a slight difference in the overall distribution of titers. The vaccinated sera exhibited a broader range of IgG titer (EC_50_ of 6×10^−5^ to 5×10^−2^) than the convalescent sera (EC_50_ of 2×10^−5^ to 8×10^−2^) against RBD-WT. Similarly, the vaccinated sera exhibited a broader heterogeneity in the titers of the RBD Delta (EC_50_ of 6×10^−5^ to 4×10^−2^) than the convalescent sera (EC_50_ of 1.5×10^−5^ to 9×10^−2^). Regarding the response to the omicron RBD variant, the vaccinated population showed significantly higher affinity than the convalescent population (EC_50_ of 1×10^−3^ and 3×10^−2^, respectively). The titer distribution of the vaccinated population was less broad and of higher titer values (lower EC_50_), showing a significant difference in the binding than that from natural immunity induced by previous infections. These findings perfectly coincide with previously reported data^36,39–42^. Finally, in two of the vaccinated patients, we found only a 2.5 to 4.5-fold lower titer of IgGs against the omicron RBD variant as compared to the RBD WT and Delta (titers IgG_P-27,28_ = 1×10^−5^ for RBD WT and Delta; IgG_P-27,28_ = 4.3×10^−4^ for Omicron).

The yeast serological assay therefore enabled this small-scale study comparing IgG titer and specificity against multiple VOCs for vaccinated and convalescent patient sera. If the sera were stratified during sampling, association studies could monitor the development of a population’s immunity against SARS-CoV-2 or virtually any pathogen with an identified target protein using our yeast test. Moreover, the yeast serological assay could offer insights into the immunity of convalescent and vaccinated populations, the effectiveness of each vaccine type, and the effects of boosters.

## Conclusion

We developed a novel serological assay based on yeast display of RBD variants that requires only 15 μL of crude serum as a sample and can be multiplexed and rapidly adapted to newly emerging RBD variant sequences. Preliminary development work on the assay demonstrated 100% specificity and 92% sensitivity on 20 serum samples from convalescent patients and 10 samples from vaccinated individuals. Our assay offers an immune profile of patients’ serum IgGs against the RBD of SARS-CoV-2 without requiring RBD antigen purification. It is rapidly adaptable for new RBD variants or other pathogens at low cost and with high precision.

Moreover, the yeast serological assay provides information about possible infection with SARS-CoV-2, and whether the patient has antibodies against a specific VOC among WT, Delta, and Omicron. It also offers insights into the immunity of convalescent and vaccinated populations, and the effect of antiCOVID-19 vaccination on IgG titers. In the future, this yeast serological setup could easily be scaled up for epidemiological population-wide studies by coupling yeast surface display, cell sorting and deep sequencing approaches to obtain a sequencing-based readout. Such an approach could allow the monitoring of multiple patient-specific immune responses to different viral epitopes simultaneously at a high throughput. We envision the yeast immunoassay as a competitive method that will find applications in various viral epidemiology or evolution projects and address current limitations in serological testing.

## Data Availability

All data produced in the present study are available upon reasonable request to the authors.

## Acknowledgments

This work was supported by the University of Basel, ETH Zurich, the Basel University Hospital, the Swiss Nanoscience Institute, the Mexican CONACYT program, the Swiss National Science Foundation (NCCR Molecular Systems Engineering), and the Botnar Research Centre for Child Health (FTC Covid-19, to MAN and YG). We thank Silke Purschke for support with clinical data handling. We thank Richard Neher for helpful communications.

