## Supplementary Information for "Rapidly Adaptable Multiplexed Yeast Surface Display Serological Assay for Immune Escape Screening of SARS-CoV-2 Variants"

**Protein Sequences**

***Yeast constructs***

- Aga2p-GS-T7-Xpresst-His6  (Empty cassette used as Negative control)

MQLLRCFSIFSVIASVLAQELTTICEQIPSPTLESTPYSLSTTTILANGKAMQGVFEYYKSVTFVSNCGSHPSTTSKGSPINTQYVFKL
LQASGGGGSGGGGSGGGGSASMTGGQQMGRDLYDDDDKVPGSVEFSTRGHHHHHH*

- Aga2p-GS-T7-Xpress-RBD_Wt-His6

MQLLRCFSIFSVIASVLAQELTTICEQIPSPTLESTPYSLSTTTILANGKAMQGVFEYYKSVTFVSNCGSHPSTTSKGSPINTQYVFKL
LQASGGGGSGGGGSGGGGSASMTGGQQMGRDLYDDDDKVPGSNITNLCPFGEVFNATRFASVYAWNRKRISNCVADYSVL
YNSASFSTFKCYGVSPTKLNDLCFTNVYADSFVIRGDEVRQIAPGQTGKIADYNYKLPDDFTGCVIAWNSNNLDSKVGGNYNYL
YRLFRKSNLKPFERDISTEIYQAGSTPCNGVEGFNCYFPLQSYGFQPTNGVGYQPYRVVVLSFELLHAPATVCGPKKSTTRGHHH
HHH*

- Aga2p-GS-T7-Xpress-RBD_Delta-His6

MQLLRCFSIFSVIASVLAQELTTICEQIPSPTLESTPYSLSTTTILANGKAMQGVFEYYKSVTFVSNCGSHPSTTSKGSPINTQYVFKL
LQASGGGGSGGGGSGGGGSASMTGGQQMGRDLYDDDDKVPGSNITNLCPFGEVFNATRFASVYAWNRKRISNCVADYSVL
YNSASFSTFKCYGVSPTKLNDLCFTNVYADSFVIRGDEVRQIAPGQTGKIADYNYKLPDDFTGCVIAWNSNNLDSKVGGNYNYR
YRLFRKSNLKPFERDISTEIYQAGSKPCNGVEGFNCYFPLQSYGFQPTNGVGYQPYRVVVLSFELLHAPATVCGPKKSTTRGHHH
HHH*

- Aga2p-GS-T7-Xpress-RBD_Omicron-His6

MQLLRCFSIFSVIASVLAQELTTICEQIPSPTLESTPYSLSTTTILANGKAMQGVFEYYKSVTFVSNCGSHPSTTSKGSPINTQYVFKL
LQASGGGGSGGGGSGGGGSASMTGGQQMGRDLYDDDDKVPGSNITNLCPFDEVFNATRFASVYAWNRKRISNCVADYSVL
YNLAPFFTFKCYGVSPTKLNDLCFTNVYADSFVIRGDEVRQIAPGQTGNIADYNYKLPDDFTGCVIAWNSNKLDSKVSGNYNYLY
RLFRKSNLKPFERDISTEIYQAGNKPCNGVAGFNCYFPLRSYSFRPTYGVGHQPYRVVVLSFELLHAPATVCGPKKSTTRGHHHH
HH*

**Table S1. List of primers and DNA sequences**

| Primer | Sequence |
|---|---|
| US_primer_F | TGACGGGTGGACAGCAAATGGGGCG |
| DS_primer_R | GGTCAGAGCACGATTGGGTCAGCGG |
| Gibson_Epi_F | CAAAGGCAGCCCCATAAACACACAGTATGTTTTTAAGCTTCTGCAGG |
| Gibson_Epi_R | GATTTTGTTACATCTACACTGTTGTTATCAGATCAGCGGGTTTAAACCTGGGTCAG |

| Plasmid | Characteristic | Source |
|---|---|---|
| pYD1 | Vector for C-terminal yeast display used as Empty cassette (Negative control) | Addgene, USA |
| pYB1 | Barcoded empty vector for C-terminal yeast display of RBD variants | This study |
| pYB1_RBD_wt | Barcoded vector for C-terminal yeast display of RBD WT | This study |
| pYB1_RBD_delta | Barcoded vector for C-terminal yeast display of RBD Delta | This study |
| pYB1_RBD_omicron | Barcoded vector for C-terminal yeast display of RBD Omicron | This study |

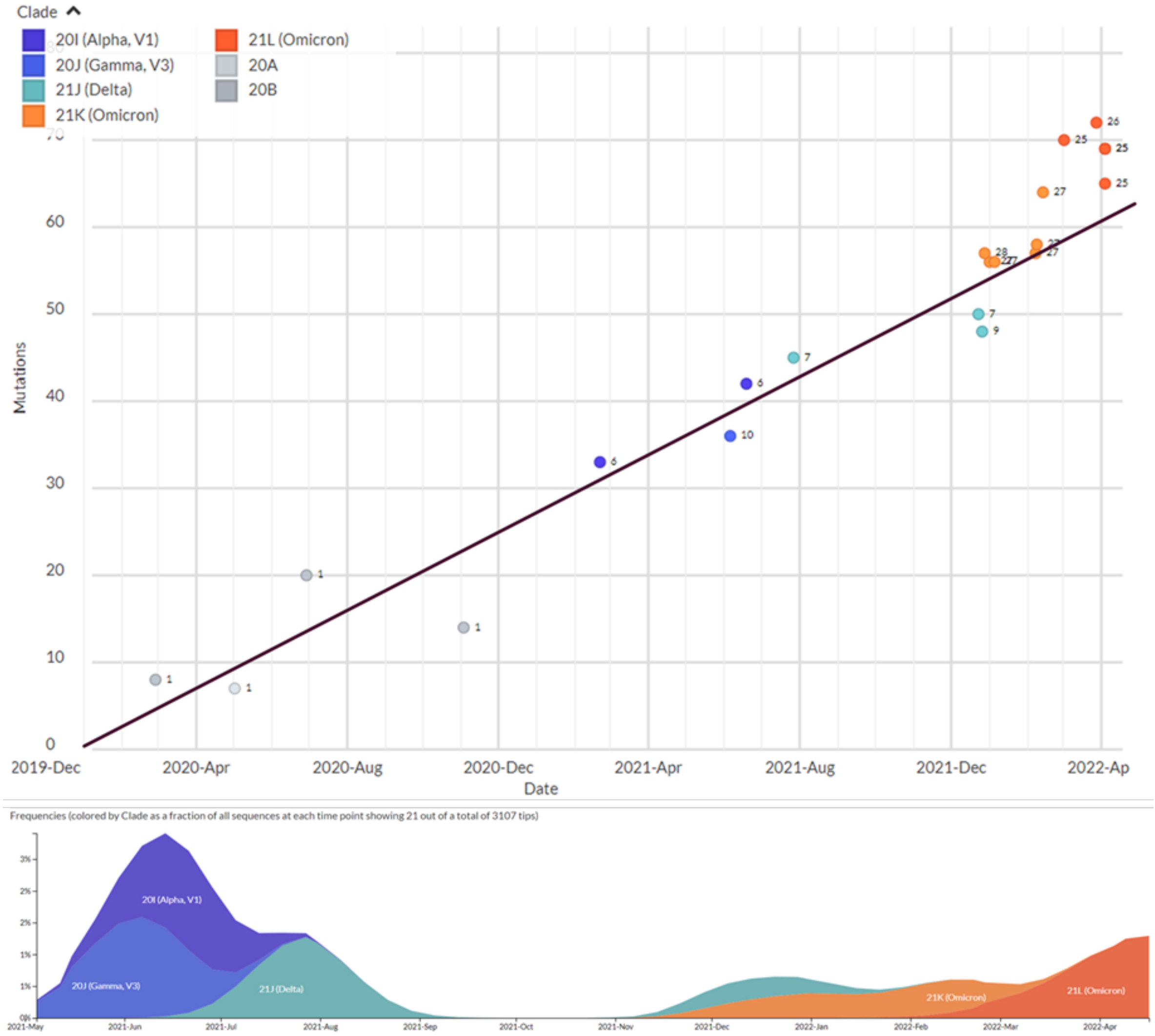


**Figure S1. Genomic epidemiology of SARS-CoV-2 with Switzerland-focused subsampling**
Upper panel: The phylogeny plot shows the evolutionary accumulation of mutations on S1 protein (harboring RBD) in Switzerland's three main SARS-CoV-2 variants of concern: Alpha, Delta, and Omicron. Approximately 15 new mutations in the Omicron RBD have been identified across Switzerland. Lower panel: The lineage frequency plot shows the emergence of new SARS-CoV-2 variant sequences over time in Switzerland. The Alpha variant overcame the Gamma in the early stages of the pandemic. The observed lower plateau corresponds to the vaccination peak and lockdown measures after the spread of the Delta outbreak. The rise and prevalence of the Omicron variant outran the vaccination and containment efforts. Time calibrated and Switzerland-focused SARS-CoV-2 phylogeny is available from the Nextstrain platform (https://nextstrain.org/ncov/gisaid/global)[43].

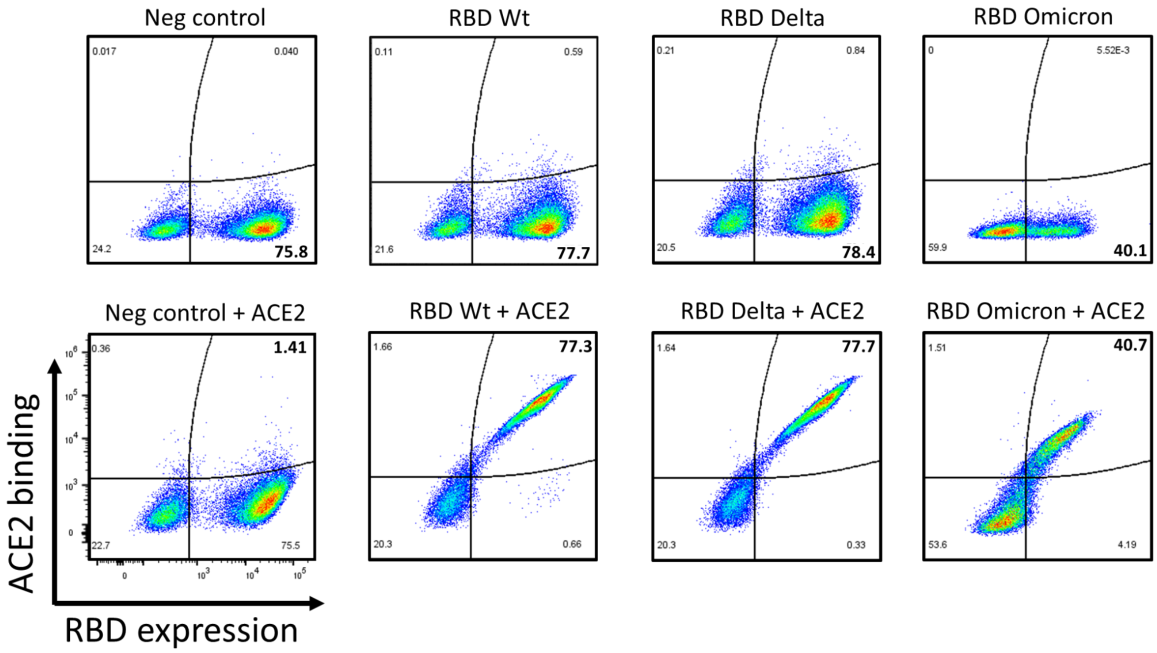


**Figure S2. Validation of ACE2 binding to RBD variants displayed on yeast cell walls**
Analytical flow cytometry of yeast displayed RBD variants binding to ACE2. Yeast cells displaying different RBD variants were labeled, and RBD display was verified by flow cytometry (upper row). A binding assay to ACE2 was performed using 200 nM of the human protein in order to verify the correct folding of the recombinant displayed constructs and the binding to ACE2.

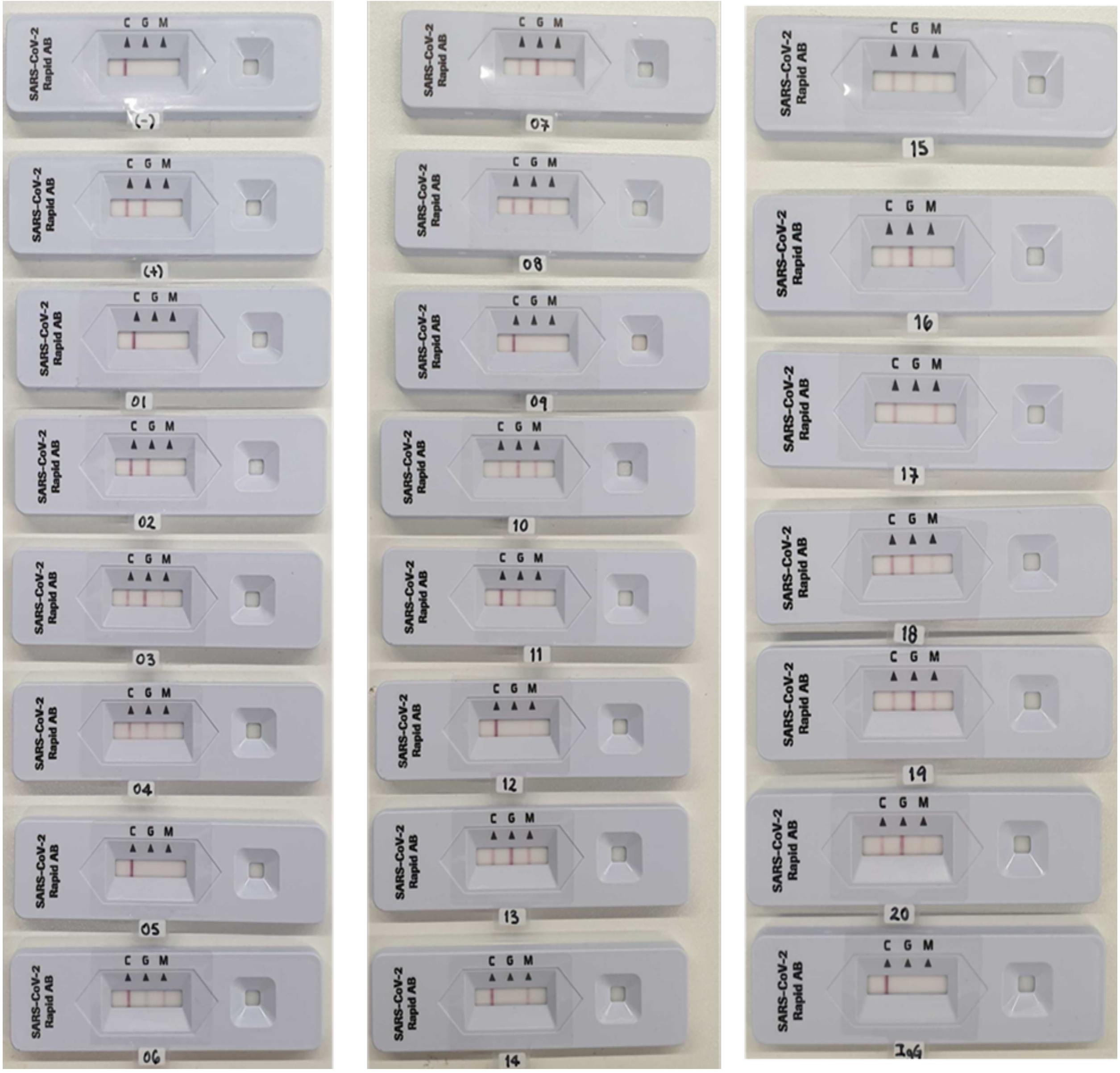


**Figure S3. Roche lateral flow test photos of convalescent sera**

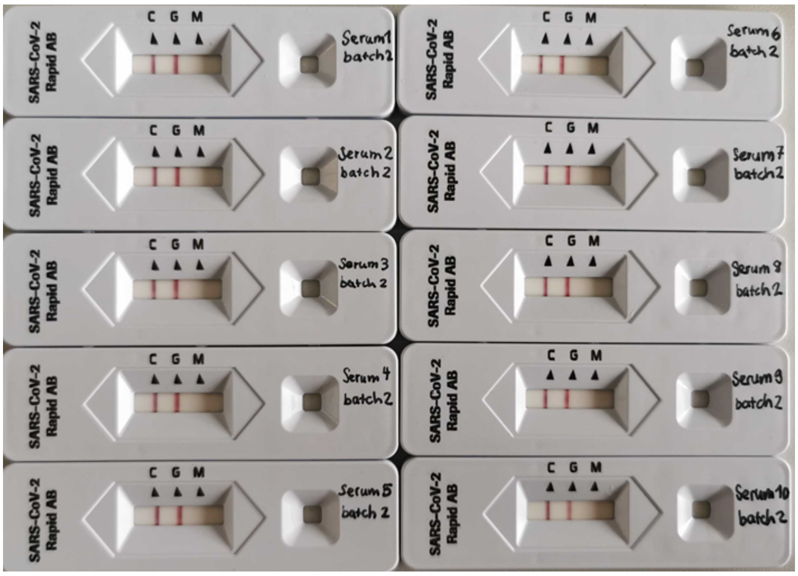

**Figure S4. Roche lateral flow test photos of vaccinated sera.**

**Table S3. On yeast serology of convalescent and vaccinated sera.**

Summary of yeast serology on patient sera. A lower $EC_{50\text{-RBD}}$ value than the $EC_{50\text{-Neg}}$ of the corresponding negative control indicates a tighter binding ($EC_{50\text{-Neg}}$ / $EC_{50\text{-RBD}}$ > 3.25 is classified as positive test result). σEC50 is the standard error of the mean (n = 2).

| Serum | RBD Variant | EC50 | σEC50 | Ratio Neg/RBD | Yeast test result | Roche test result |
|---|---|---|---|---|---|---|
| 1 | Neg ctrl | 9.95E-03 | 6.75E-04 | 1.00 | - | (-) |
| | WT | 1.18E-02 | 4.25E-04 | 0.85 | (-) | |
| | Delta | 1.62E-02 | 1.05E-03 | 0.61 | (-) | |
| | Omicron | 2.16E-02 | 8.78E+00 | 0.46 | (-) | |
| 2 | Neg ctrl | 2.30E-03 | 1.11E-04 | 1.00 | - | (+) |
| | WT | 2.74E-03 | 1.07E-04 | 0.84 | (-) | |
| | Delta | 2.71E-03 | 1.16E-04 | 0.85 | (-) | |
| | Omicron | 9.46E-03 | 5.97E-04 | 0.24 | (-) | |
| 3 | Neg ctrl | 2.40E-03 | 9.60E-05 | 1.00 | - | (+) |
| | WT | 3.85E-04 | 8.00E-06 | 6.23 | (+) | |
| | Delta | 3.83E-04 | 1.30E-05 | 6.26 | (+) | |
| | Omicron | 3.96E-03 | 1.07E-04 | 0.60 | (-) | |
| 4 | Neg ctrl | 5.56E-03 | 4.07E-04 | 1.00 | - | (+) |
| | WT | 3.02E-04 | 8.00E-06 | 18.41 | (+) | |
| | Delta | 3.18E-04 | 1.30E-05 | 17.49 | (+) | |
| | Omicron | 6.52E-03 | 2.36E-04 | 0.85 | (-) | |
| 5 | Neg ctrl | 9.44E-04 | 6.90E-05 | 1.00 | - | (-) |
| | WT | 1.60E-03 | 1.02E-04 | 0.59 | (-) | |
| | Delta | 1.39E-03 | 1.36E-04 | 0.68 | (-) | |
| | Omicron | 2.34E-03 | 9.80E-05 | 0.40 | (-) | |
| 6 | Neg ctrl | 2.11E-03 | 1.59E-04 | 1.00 | - | (+) |
| | WT | 1.62E-04 | 7.00E-06 | 13.01 | (+) | |
| | Delta | 1.94E-04 | 4.00E-06 | 10.87 | (+) | |
| | Omicron | 5.33E-03 | 2.91E-04 | 0.40 | (-) | |
| 7 | Neg ctrl | 9.89E-04 | 8.30E-05 | 1.00 | - | (+) |
| | WT | 1.16E-04 | 3.00E-06 | 8.53 | (+) | |
| | Delta | 1.39E-04 | 4.00E-06 | 7.12 | (+) | |
| | Omicron | 1.37E-03 | 5.40E-05 | 0.72 | (-) | |
| 8 | Neg ctrl | 4.24E-02 | 1.12E-01 | 1.00 | - | (+) |
| | WT | 1.01E-04 | 5.00E-06 | 419.84 | (+) | |
| | Delta | 1.10E-04 | 6.00E-06 | 385.49 | (+) | |
| | Omicron | 2.96E-03 | 1.99E-04 | 14.33 | (+) | |
| 9 | Neg ctrl | 6.01E-04 | 9.40E-05 | 1.00 | - | (-) |
| | WT | 7.87E-04 | 3.70E-05 | 0.76 | (-) | |
| | Delta | 9.64E-04 | 5.10E-05 | 0.62 | (-) | |
| | Omicron | 1.38E-03 | 4.80E-05 | 0.44 | (-) | |
| 10 | Neg ctrl | 9.28E-04 | 4.51E-04 | 1.00 | - | (+) |
| | WT | 1.80E-05 | 1.00E-06 | 51.56 | (+) | |
| | Delta | 2.20E-05 | 2.00E-06 | 42.18 | (+) | |
| | Omicron | 6.22E-04 | 2.90E-05 | 1.49 | (-) | |

**Table S3 (continued). On yeast serology of convalescent and vaccinated sera.**

| Serum | RBD Variant | EC50 | σEC50 | Ratio Neg/RBD | Yeast test result | Roche test result |
|---|---|---|---|---|---|---|
| 11 | Neg ctrl | 2.16E-03 | 3.80E-05 | 1.00 | - | (+) |
| | WT | 6.18E-03 | 3.19E-04 | 0.35 | (-) | |
| | Delta | 6.99E-03 | 3.74E-04 | 0.31 | (-) | |
| | Omicron | 8.85E-03 | 3.64E-04 | 0.24 | (-) | |
| 12 | Neg ctrl | 3.13E-03 | 3.73E-04 | 1.00 | - | (-) |
| | WT | 3.31E-03 | 8.00E-06 | 0.94 | (-) | |
| | Delta | 3.55E-03 | 2.54E-04 | 0.88 | (-) | |
| | Omicron | 6.38E-03 | 4.02E-04 | 0.49 | (-) | |
| 13 | Neg ctrl | 7.50E-04 | 2.84E-04 | 1.00 | - | (+) |
| | WT | 6.30E-05 | 2.00E-06 | 11.90 | (+) | |
| | Delta | 5.70E-05 | 2.00E-06 | 13.16 | (+) | |
| | Omicron | 1.67E-03 | 1.91E-04 | 0.45 | (-) | |
| 14 | Neg ctrl | 6.66E-04 | 3.30E-05 | 1.00 | - | (+) |
| | WT | 1.52E-03 | 9.70E-05 | 0.44 | (-) | |
| | Delta | 1.89E-03 | 1.35E-04 | 0.35 | (-) | |
| | Omicron | 4.19E-03 | 2.38E-04 | 0.16 | (-) | |
| 15 | Neg ctrl | 2.85E-03 | 1.42E-04 | 1.00 | - | (+) |
| | WT | 2.80E-04 | 8.00E-06 | 10.17 | (+) | |
| | Delta | 2.52E-04 | 1.20E-05 | 11.30 | (+) | |
| | Omicron | 4.35E-03 | 3.07E-04 | 0.65 | (-) | |
| 16 | Neg ctrl | 8.82E-03 | 1.19E-03 | 1.00 | - | (+) |
| | WT | 3.50E-05 | 2.00E-06 | 251.89 | (+) | |
| | Delta | 3.50E-05 | 2.00E-06 | 251.89 | (+) | |
| | Omicron | 1.42E-03 | 9.60E-05 | 6.23 | (+) | |
| 17 | Neg ctrl | 2.50E-03 | 2.28E-04 | 1.00 | - | (+) |
| | WT | 1.09E-03 | 6.20E-05 | 2.29 | (-) | |
| | Delta | 1.28E-03 | 7.80E-05 | 1.95 | (-) | |
| | Omicron | 3.83E-03 | 1.61E-04 | 0.65 | (-) | |
| 18 | Neg ctrl | 6.19E-03 | 5.90E-04 | 1.00 | - | (+) |
| | WT | 8.40E-05 | 3.00E-06 | 73.67 | (+) | |
| | Delta | 6.60E-05 | 4.00E-06 | 93.76 | (+) | |
| | Omicron | 2.78E-03 | 1.35E-04 | 2.23 | (-) | |
| 19 | Neg ctrl | 2.63E-03 | 2.60E-04 | 1.00 | - | (+) |
| | WT | 4.97E-04 | 2.30E-05 | 5.28 | (+) | |
| | Delta | 4.94E-04 | 1.60E-05 | 5.31 | (+) | |
| | Omicron | 3.73E-03 | 3.93E-04 | 0.70 | (-) | |
| 20 | Neg ctrl | 6.86E-04 | 7.30E-05 | 1.00 | - | (+) |
| | WT | 5.30E-05 | 3.00E-06 | 12.94 | (+) | |
| | Delta | 7.00E-05 | 3.00E-06 | 9.80 | (+) | |
| | Omicron | 8.32E-04 | 5.30E-05 | 0.82 | (-) | |

**Table S3 (continued). On yeast serology of convalescent and vaccinated sera.**

| Serum | RBD Variant | EC50 | σEC50 | Ratio Neg/RBD | Yeast test result | Roche test result |
|---|---|---|---|---|---|---|
| 21 | Neg ctrl | 1.36E-03 | 6.40E-05 | 1.00 | - | (+) |
| | WT | 1.06E-04 | 3.00E-06 | 12.80 | (+) | |
| | Delta | 1.41E-04 | 4.00E-06 | 9.62 | (+) | |
| | Omicron | 2.03E-03 | 6.30E-05 | 0.67 | (-) | |
| 22 | Neg ctrl | 6.92E-03 | 3.98E-04 | 1.00 | - | (+) |
| | WT | 2.00E-05 | 1.00E-06 | 346.00 | (+) | |
| | Delta | 2.60E-05 | 2.00E-06 | 266.15 | (+) | |
| | Omicron | 4.26E-04 | 2.40E-05 | 16.24 | (+) | |
| 23 | Neg ctrl | 4.98E-03 | 2.26E-04 | 1.00 | - | (+) |
| | WT | 3.00E-05 | 1.00E-06 | 165.97 | (+) | |
| | Delta | 2.90E-05 | 1.00E-06 | 171.69 | (+) | |
| | Omicron | 5.80E-04 | 2.60E-05 | 8.58 | (+) | |
| 24 | Neg ctrl | 2.16E-02 | 2.88E-02 | 1.00 | - | (+) |
| | WT | 1.76E-04 | 5.00E-06 | 122.53 | (+) | |
| | Delta | 2.14E-04 | 7.00E-06 | 100.77 | (+) | |
| | Omicron | 2.60E-03 | 1.39E-04 | 8.31 | (+) | |
| 25 | Neg ctrl | 1.34E-03 | 9.50E-05 | 1.00 | - | (+) |
| | WT | 6.20E-05 | 2.00E-06 | 21.58 | (+) | |
| | Delta | 8.10E-05 | 3.00E-06 | 16.52 | (+) | |
| | Omicron | 1.40E-03 | 8.00E-05 | 0.96 | (-) | |
| 26 | Neg ctrl | 5.24E-03 | 3.73E-04 | 1.00 | - | (+) |
| | WT | 8.80E-05 | 2.00E-06 | 59.51 | (+) | |
| | Delta | 8.60E-05 | 2.00E-06 | 60.90 | (+) | |
| | Omicron | 1.01E-03 | 2.60E-05 | 5.18 | (+) | |
| 27 | Neg ctrl | 6.39E-03 | 2.83E-04 | 1.00 | - | (+) |
| | WT | 1.00E-05 | 0.00E+00 | 639.20 | (+) | |
| | Delta | 1.00E-05 | 0.00E+00 | 639.20 | (+) | |
| | Omicron | 4.33E-04 | 1.60E-05 | 14.76 | (+) | |
| 28 | Neg ctrl | 2.00E-02 | 2.31E-04 | 1.00 | - | (+) |
| | WT | 1.90E-05 | 2.00E-06 | 1051.26 | (+) | |
| | Delta | 1.70E-05 | 2.00E-06 | 1174.94 | (+) | |
| | Omicron | 4.24E-04 | 3.10E-05 | 47.11 | (+) | |
| 29 | Neg ctrl | 2.36E-03 | 1.52E-04 | 1.00 | - | (+) |
| | WT | 8.90E-05 | 1.40E-05 | 26.46 | (+) | |
| | Delta | 8.10E-05 | 1.00E-06 | 29.07 | (+) | |
| | Omicron | 8.59E-04 | 2.20E-05 | 2.74 | (-) | |
| 30 | Neg ctrl | 2.51E-03 | 7.00E-05 | 1.00 | - | (+) |
| | WT | 6.71E-04 | 1.90E-05 | 3.75 | (+) | |
| | Delta | 7.74E-04 | 5.50E-05 | 3.25 | (+) | |
| | Omicron | 1.69E-03 | 7.10E-05 | 1.49 | (-) | |

**Table S3 (continued). On yeast serology of commercial SARS-Cov-2 Negative sera.**
Summary of yeast serology on validated single-donor human SARS-Cov-2-negative control sera (Innovative Research, USA; Cat: ISERSCOV2N100UL).

| Serum | RBD Variant | EC50 | σEC50 | Ratio Neg/RBD | Yeast test result | Roche test result |
|---|---|---|---|---|---|---|
| 31 | Neg ctrl | 8.60E-05 | 3.00E-06 | 1.00 | - | (-) |
| | WT | 5.54E-04 | 9.00E-06 | 0.16 | (-) | |
| | Delta | 4.87E-04 | 8.00E-06 | 0.18 | (-) | |
| | Omicron | 9.72E-04 | 2.90E-05 | 0.09 | (-) | |
| 32 | Neg ctrl | 1.13E-02 | 4.24E-04 | 1.00 | - | (-) |
| | WT | 1.07E-02 | 7.12E-04 | 1.06 | (-) | |
| | Delta | 1.02E-02 | 8.76E-04 | 1.11 | (-) | |
| | Omicron | 1.09E-02 | 1.23E-03 | 1.04 | (-) | |
| 33 | Neg ctrl | 4.88E-03 | 1.88E-04 | 1.00 | - | (-) |
| | WT | 3.88E-03 | 1.05E-04 | 1.26 | (-) | |
| | Delta | 3.72E-03 | 1.41E-04 | 1.31 | (-) | |
| | Omicron | 3.72E-03 | 1.24E-04 | 1.31 | (-) | |
| 34 | Neg ctrl | 7.34E-03 | 2.00E-04 | 1.00 | - | (-) |
| | WT | 6.38E-03 | 3.21E-04 | 1.15 | (-) | |
| | Delta | 7.11E-03 | 3.85E-04 | 1.03 | (-) | |
| | Omicron | 7.75E-03 | 4.24E-04 | 0.95 | (-) | |
| 35 | Neg ctrl | 7.65E-03 | 2.06E-04 | 1.00 | - | (-) |
| | WT | 9.05E-03 | 4.58E-04 | 0.85 | (-) | |
| | Delta | 7.57E-03 | 2.06E-04 | 1.01 | (-) | |
| | Omicron | 8.79E-03 | 4.26E-04 | 0.87 | (-) | |

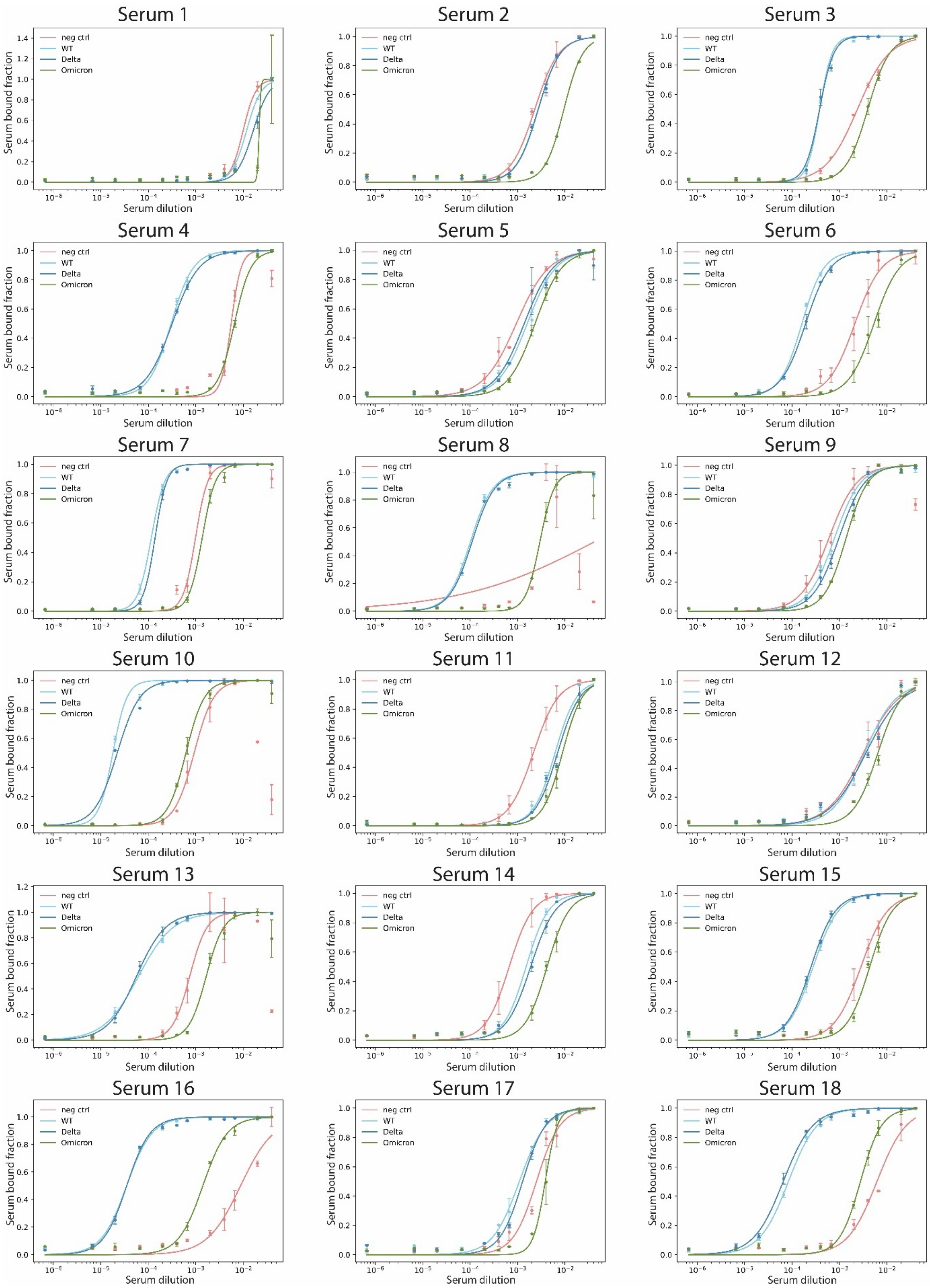


**Figure S5. Titration curves of convalescent sera.**

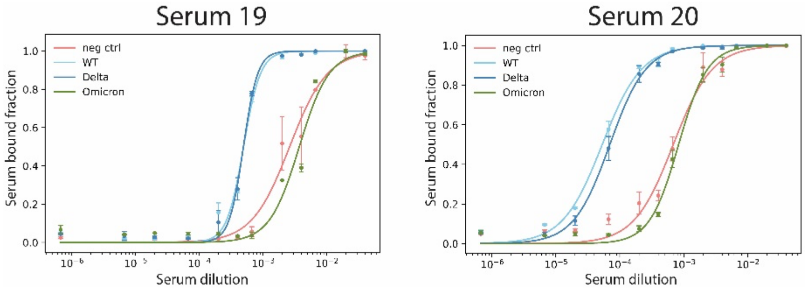


**Figure S5 (continued). Titration curves of convalescent sera.**

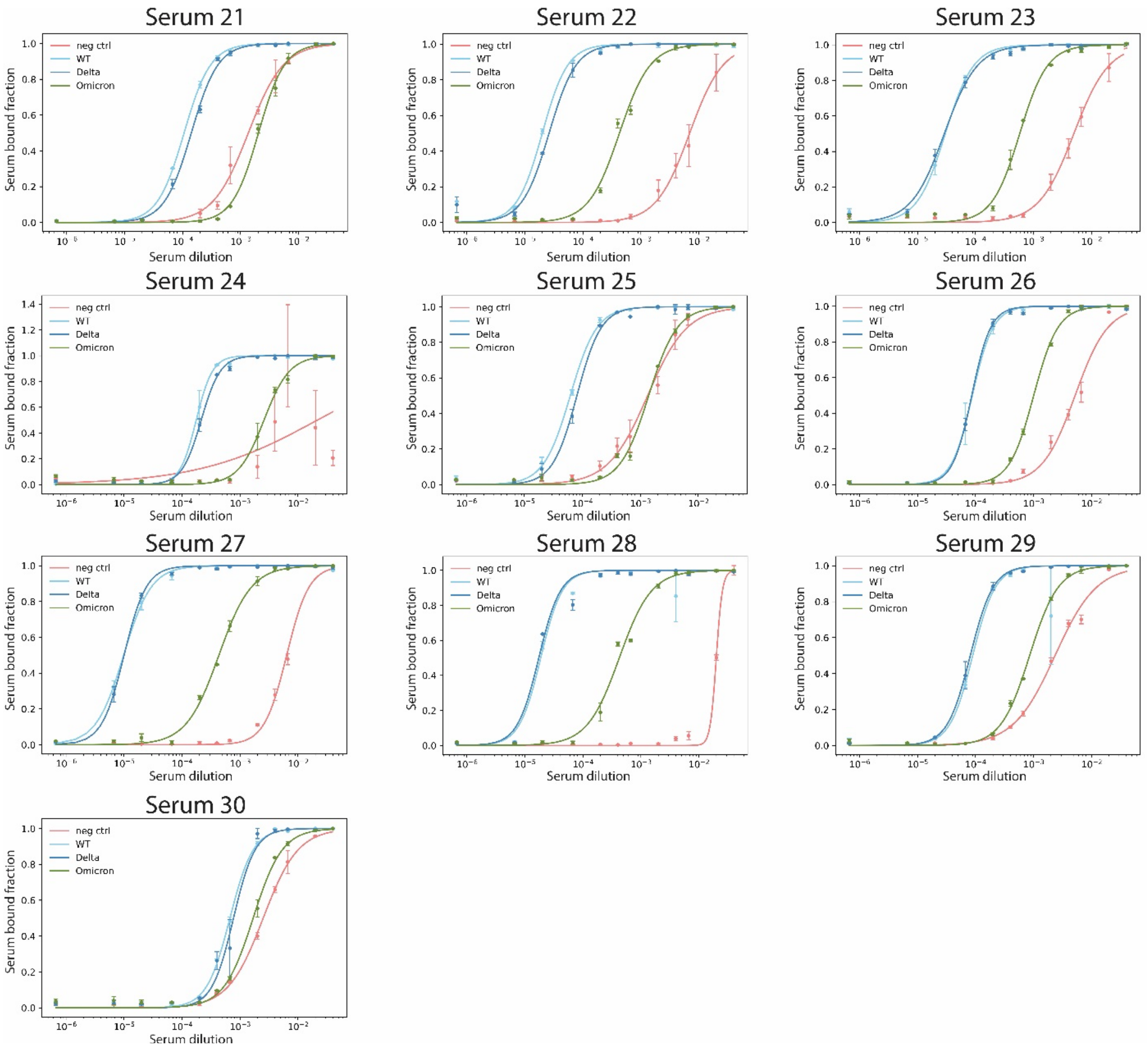


**Figure S6. Titration curves of vaccinated sera.**

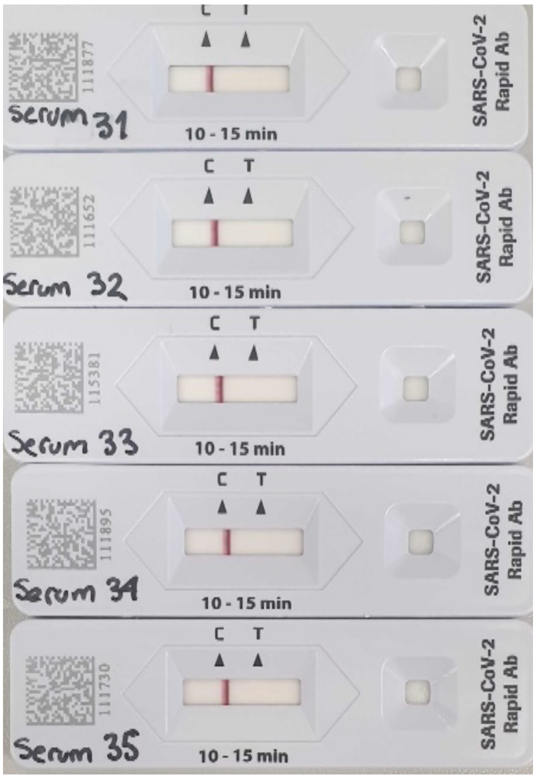


**Figure S7. Roche lateral flow test photos of negative control sera**

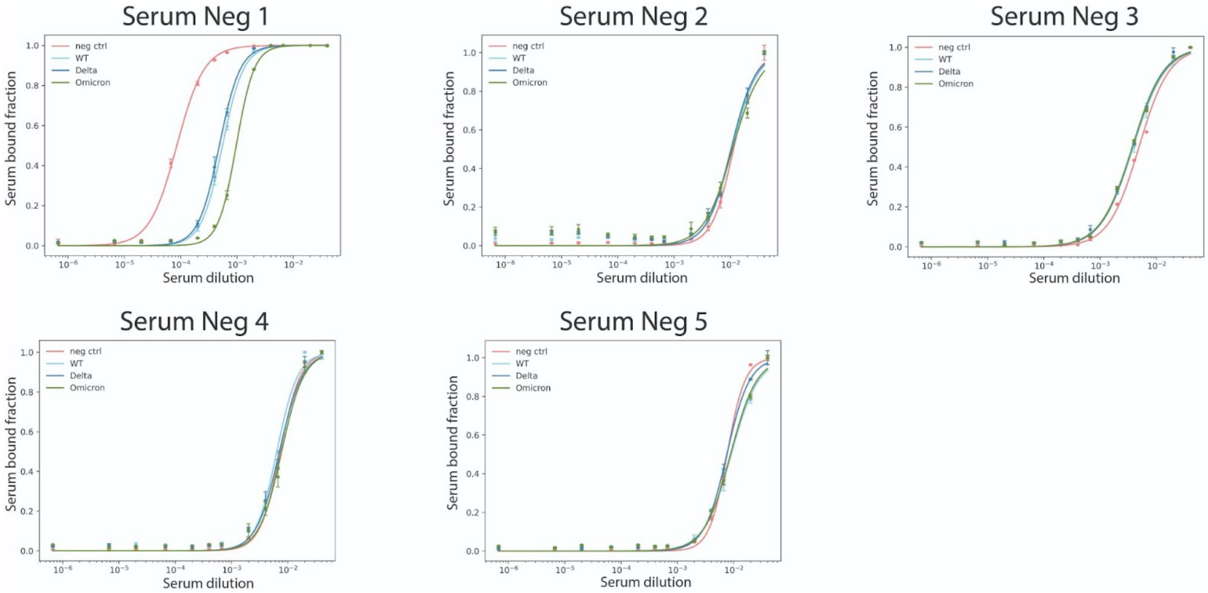


**Fig S8. Titration curves of negative control sera.** These samples were numbered as 31-35 in Table S3.

| | | Reference test | | |
|---|---|---|---|---|
| | | Roche (+) | Roche (-) | |
| Index test | Yeast (+) | 22 | 0 | 22 |
| | Yeast (-) | 2 | 10 | 12 |
| | | 24 | 10 | 34 |
| | **Sens=** | 0.92 | **PPV=** | 1 |
| | **Spec=** | 1 | **NPV=** | 0.83 |

**Fig S9. Confusion matrix for the yeast SARS-CoV-2 RBD WT serology test**

The table was built using the frequency yeast immunoassay (index test) results for only SARS-CoV-2 RBD WT because the reference test (Roche SARS-CoV-2 IgG/IgM rapid antibody lateral flow) cannot differentiate among RBD variants. Accuracy metrics are shown below the confusion matrix. Sens: sensitivity; Spec: specificity; PPV: positive predictive value; NPV: negative predictive value.

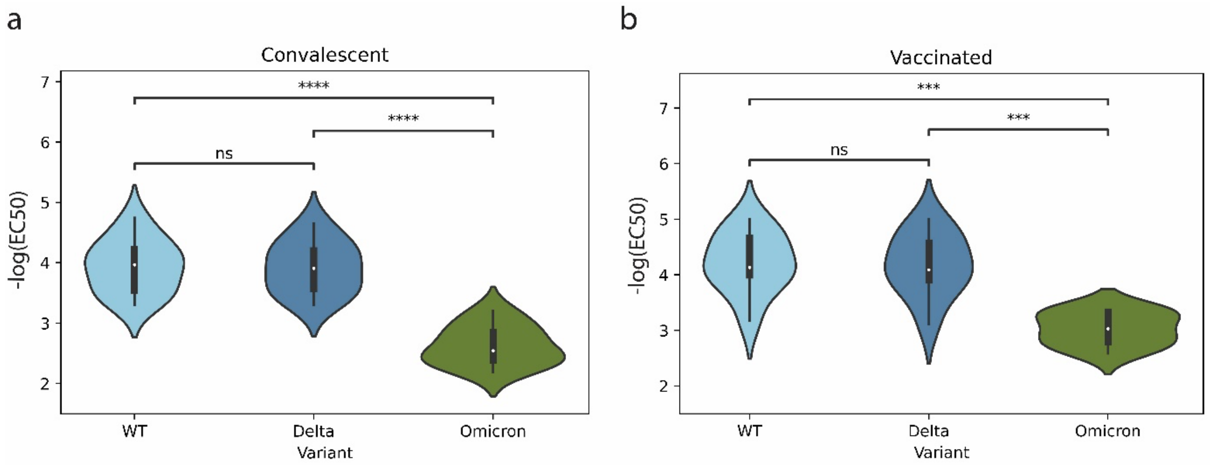


**Figure S10. Comparative analysis of immunity between convalescent and vaccinated sera**

**a.** Comparison of IgG titers against different RBD variants among convalescent sera. **b.** Comparison of IgG titers against different RBD variants among vaccinated sera. Significance from ANOVA is shown as n.s.: $p \geq 1.0$; **: $1.0\text{x}10^{-3} < p \leq 1.0\text{x}10^{-2}$; ***: $1.0\text{x}10^{-4} < p \leq 1.0\text{x}10^{-3}$; ****: $p \leq 1.0\text{x}10^{-4}$.
